# Dynamic Data-Driven Algorithm to Predict the Cumulative COVID-19 Infected Cases Using Susceptible-Infected-Susceptible Model

**DOI:** 10.1101/2021.03.24.21253599

**Authors:** Abhinav Anand, Saurabh Kumar, Palash Ghosh

## Abstract

In recent times, researchers have used Susceptible-Infected-Susceptible (SIS) model to understand the spread of pandemic COVID-19. The SIS model has two compartments, susceptible and infected. In this model, the interest is to determine the number of infected people at a given time point. However, it is also essential to know the cumulative number of infected people at a given time point, which is not directly available from the SIS model’s present structure. In this work, we propose a modified structure of the SIS model to determine the cumulative number of infected people at a given time point. We develop a dynamic data-driven algorithm to estimate the model parameters based on an optimally chosen training phase to predict the same. We demonstrate the proposed algorithm’s prediction performance using COVID-19 data from Delhi, India’s capital city.

## I. Introduction

The use of epidemiological models to control the spread of disease and predict the course of an outbreak has a long history. In 1760, Daniel Bernoulli proposed a mathematical model for smallpox [1]. At the beginning of the 20^*th*^ century, William Hamer and Ronald Ross studied the epidemic behavior using the law of mass action [2], [3]. In recent times, the use of epidemiological models is inevitable for better management of an infectious disease.

We have seen the use of various epidemiological models to combat the recent outbreak of Coronavirus disease 2019 (COVID-19). COVID-19 was first reported in Wuhan city of China but soon spread to other parts of the world [4]. Many authors have used some version of Susceptible-Infected-Recovered (SIR) models to predict the COVID-19 outbreak in different countries or regions [5]–[7]. The basic SIR model assumes that the infected individuals are either recovered (and immune) from the disease or died [8]. It also assumes the number of deaths from the disease is negligible compared to the total population. However, WHO mentioned that “there is currently no evidence that people who have recovered from COVID-19 and have antibodies are protected from a second infection” [9]. For example, health authorities in South Korea noticed that 163 patients became COVID-19 positive again after a full recovery [10]. Several studies have found that individuals who are infected by the COVID-19 may build short-term immunity against the disease, and there is no long-lasting guaranteed protection [11]–[13]. In this context, when there is no long-term protection from the disease after infection, the Susceptible-Infected-Susceptible (SIS) model is appropriate. In an SIS model, people who recover from the disease are added to the susceptible compartment as they can be infected again. In this work, we consider the SIS model to predict the COVID-19 outbreak.

In an SIS model, the main focus is to determine the number of infected people at a given time point. However, it is also essential for planning purposes to know the cumulative number of infected people at a given time point. One cannot directly find the cumulative number of infected people from the SIS model’s present structure. In this work, our main contribution is to provide an SIS model-structure which can give the cumulative number of infected people easily. We incorporate a death due to disease compartment in the SIS model to estimate the model parameters accurately. We develop a dynamic data-driven algorithm to estimate the model parameters efficiently to predict the cumulative infected cases. In this process, we show how to select the optimal training phase to build the model. Finally, the developed algorithm has been implemented using COVID-19 data from Delhi, India’s capital city. We also provide an R-package so that users can easily implement the developed model with their data.

## II. Susceptible-Infected-Susceptible (SIS) model

In an SIS model [14], there are only two compartments, Susceptible and Infected. An SIS model assumes that an individual has not developed any long-term immune against the disease after infection and thus is at risk of re-infection; hence, it gets added back to the susceptible population. In other words, as shown in Figure 1, after recovering from an infection, an individual again becomes susceptible. Examples of such infections are the common cold and influenza.

**Fig. 1.**
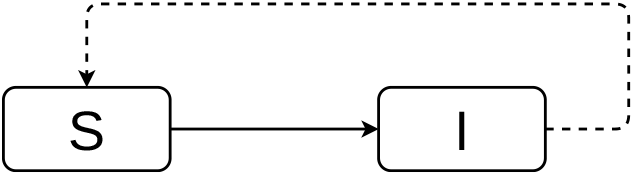
Pictorial representation of an SIS Model, where ***S*** stands for susceptible and ***I*** for infected

These equations can well describe an SIS model [8],

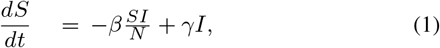

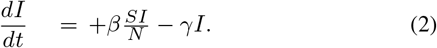

Here *t* denotes time. In this work, a day is the smallest unit of time *t*. However, one can choose other suitable units as necessary. *S* and *I* are the susceptible and the infected number of people in the population, respectively. The total population size is *N*, which is the sum of susceptible (*S*) and infected (*I*) populations. The parameter *β*, transmission rate, is the product of the contact rates among infected, and transmission probability [8], [15]. In other words, the parameter *β* is the average number of individuals infected per unit time (a day) from an infected person. Here, by assumption, *I* infected individuals can contact some individuals randomly; a fraction of *S/N* of them will be susceptible. The parameter *γ* is the recovery rate. It is assumed to be 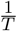 where *T* is the average duration for which infection lasts in an infected person [16], [17]. Equation (1) shows the rate of change in the susceptible (*S*) population; whereas equation (2) depicts the rate of change in the infected (*I*) population. The term 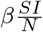 denotes the number of susceptible people infected daily and is removed from the susceptible compartment and added to the infected compartment.

The *γI* denotes the number of people recovered daily and is added back to the susceptible compartment and removed from the infected compartment. Figure 2 shows a simulated SIS model with *S* +*I* = *N* at all time points. Notice that one cannot get the cumulative infected cases directly from the above SIS model. The death due to the disease is not adjusted into this model. It may affect the efficiency of estimating model parameters when the number of deaths due to disease is not negligible (as observed in COVID-19). In the next section, we consider a modified SIS model to address these two issues.

**Fig. 2.**
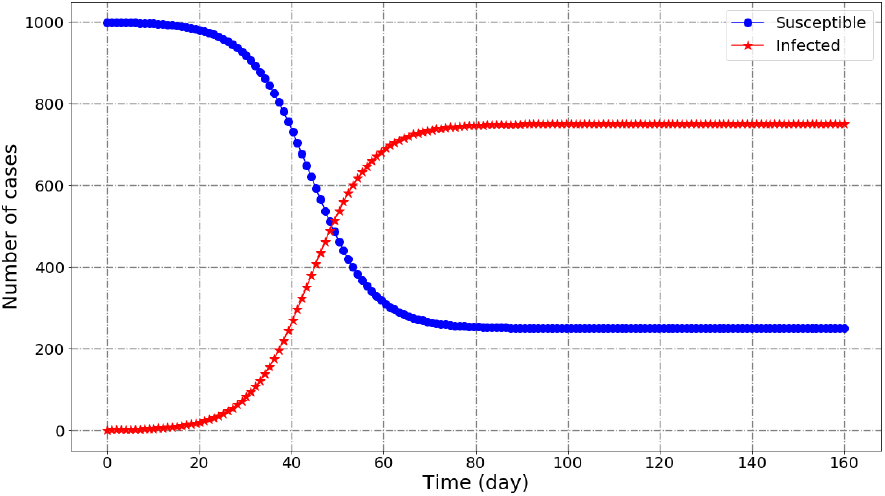
A simulated SIS model with the initial values of ***S, I*** as ***S*(0) = 1000** and ***I*(0) = 1**, respectively. Here, ***β* = 0**.**2** and ***γ* = 0**.**05**.

### A. Model Equations for modified SIS model

The proposed model can be well described in these equations,

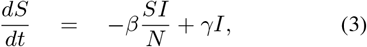

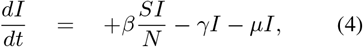

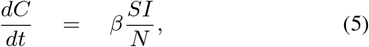

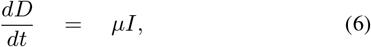

where *S, I, N, β* and *γ* are the same as defined earlier. The *C* is the cumulative infected cases from the beginning. It includes every person who is infected or was infected. The *D* is the deceased population due to the disease. Note that *D* does not include death counts from other causes. We assume that the death rate from other causes not involving the concerned disease is the same as the birth rate.

Equation (3) is the same as (1). Equation (4) represents the effective change in the infected compartment. As explained earlier, 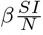 number of susceptible individuals get infected daily and are added to this compartment. The *µ* is the mortality rate of the infection. Thus, *γI* number of infected individuals are recovered from infection daily, and *µI* number of infected cases are fatal daily, are removed from this compartment. Equation (5) represents the rate of change in the cumulative infected cases, which is equal to daily infected cases 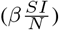. Equation (6) represents the rate of change in the deceased compartment which is equal to *µI*.

## III. Dynamic Data-Driven Algorithm to Estimate the *β*

In general, the SIS model parameters are constant for the entire duration of the study period. When the disease under consideration is present in the community for a longer time, the estimated parameter based on the entire study period may not give the right picture. For example, the COVID-19 disease outspread is highly unpredictable in the long term because the contact rates and transmission probabilities are changing over time. They vary due to various reasons like control measures implemented by respective governments. Therefore, it may be appropriate to train an SIS model with a shorter training phase and make short-term predictions. Here, the ‘*dynamic data-driven algorithm*’ means the training phase, used to estimate the model parameters, is dynamic (not fixed) and optimally chosen based on the appropriate historical data.

The two phases of the study period are the training and the prediction phases. Figure 3 shows how the study period is divided into different parts for estimation purpose. We define the four-time variables as follows:

**Fig. 3.**
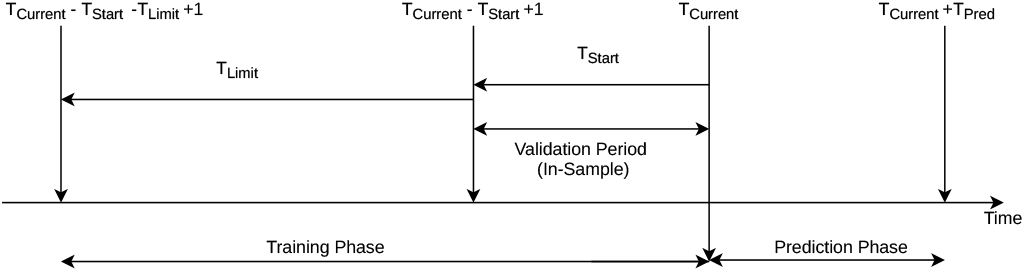
Different time points of training and prediction phases.

- *T*_*Current*_: Denotes the *date* when the training phase ends. After this date, the prediction phase starts.
- *T*_*Start*_: Denotes the minimum length of the training phase (in days). The minimum training phase is the interval, [*T*_*Current*_ *− T*_*Start*_ + 1, *T*_*Current*_].
- *T*_*Pred*_: Denotes the length of the prediction phase (in days). The prediction phase is the interval, [*T*_*Current*_ +1, *T*_*Current*_ + *T*_*Pred*_].
- *T*_*Limit*_: Denotes the upper limit of the number of additional days that can be added to the minimum training phase to optimally choose the training phase. The length of the training phase keeps increasing with a step of 1 day. Therefore, the maximum training phase interval can be, [*T*_*Current*_ *− T*_*Start*_ *− T*_*Limit*_ + 1, *T*_*Current*_].

Here, our objective is to choose an optimal training phase that can be used to predict the near future accurately. Note that for each value of 0 ≤ *t* ≤ *T*_*Limit*_, there is a different training phase denoted by [*T*_*Current*_ *− T*_*Start*_ + 1*− t, T*_*Current*_]. The minimum training phase, for *t* = 0, is [*T*_*Current*_ *− T*_*Start*_ + 1, *T*_*Current*_], whereas the maximum training phase, for *t* = *T*_*Limit*_, is [*T*_*Current*_ *−T*_*Start*_ *− T*_*Limit*_ + 1, *T*_*Current*_]. We consider the minimum training phase as the fixed in-sample validation period to compare different models based on different training phases. Here, ‘in-sample’ refers that the validation period is a subset of the considered training phase. The optimal criteria to choose the appropriate training phase is defined by the root mean squared error,

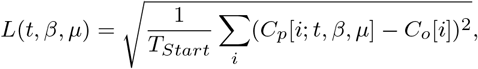

where *T*_*Current*_ *— T*_*Start*_ + 1 ≤ *i* ≤ *T*_*Current*_ ; *C*_*o*_[*i*] denotes the observed cumulative infected cases on *i*^*th*^ day and *C*_*p*_[*i*; *t, β, µ*] denotes the predicted cumulative infected cases on *i*^*th*^ day considering [*T*_*Current*_*— T*_*Start*_ + 1 *− t, T*_*Current*_] as the training phase. The optimal value of *t* is obtained as

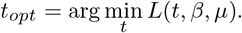

Finally, we obtain the optimal training phase that can be used for future prediction as [*T*_*Current*_ *− T*_*Start*_ + 1 *− t*_*opt*_, *T*_*Current*_].

There are three parameters in the modified SIS model, namely, *β, γ*, and *µ*. As argued earlier, 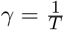, where *T* is the average duration for which infection lasts in an infected person. In case of COVID-19, *T* is taken as 14 [16], [17]. Given a training phase, Algorithm 1 is used to find the estimates of *β* and *µ* by minimizing *L*(*t, β, µ*).

### Algorithm 1: Dynamic Data-Driven Algorithm to Estimate the *β* and *µ*

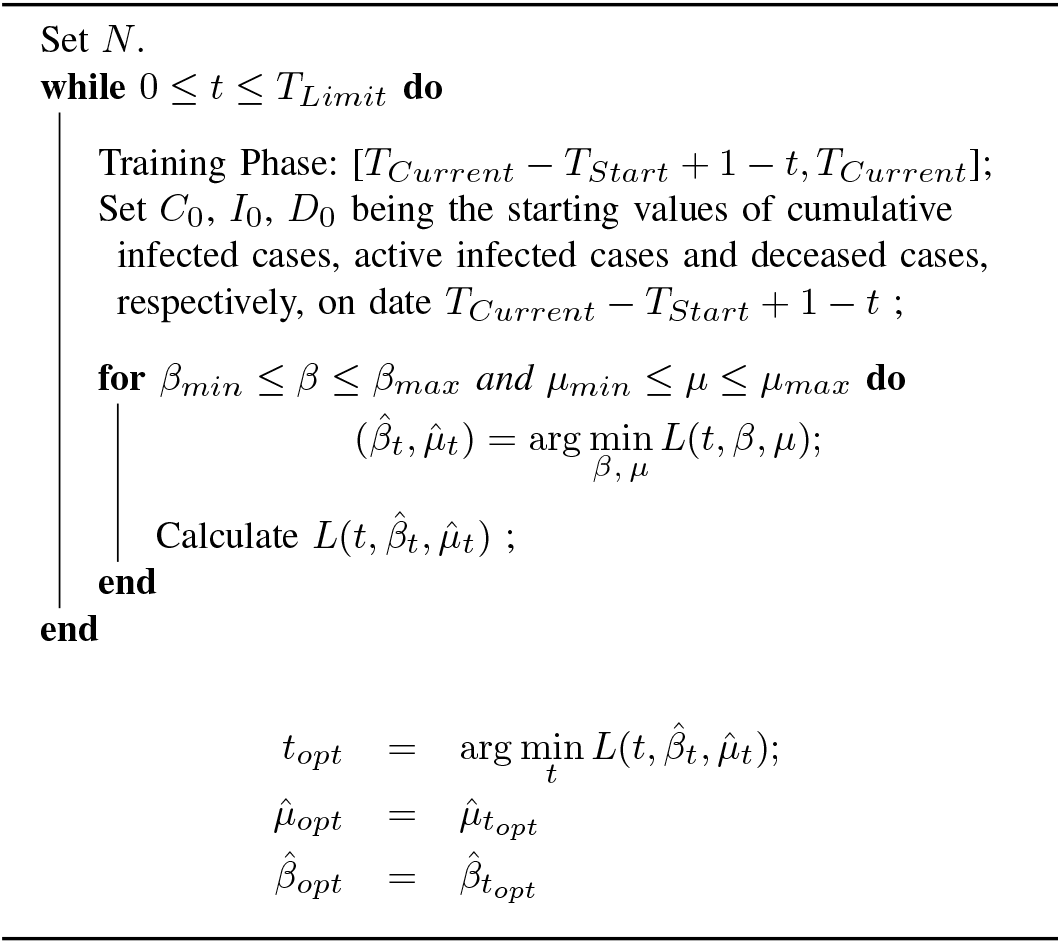

### Algorithm 2: Prediction Algorithm for the Cumulative Infected cases (*C*)

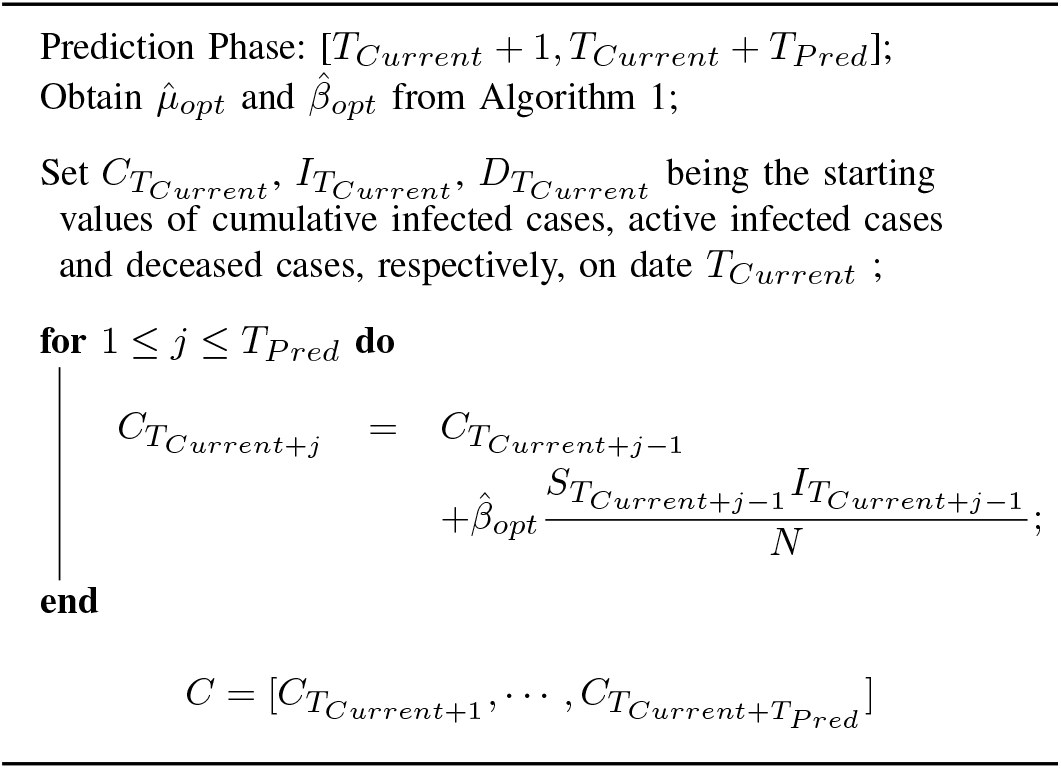

## IV. Prediction of cumulative infected cases

The 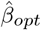 and 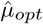 are the optimum values of *β* and *µ*, respectively, using Algorithm 1. Using Algorithm 2, the predicted values of the cumulative infected cases (*C*) are obtained for every day starting from *T*_*Current*+1_ to *T*_*Current*+*T*__*Pred*_. Here, the prediction period’s length depends on the user-supplied value of *T*_*Pred*_. For 1 ≤ *i* ≤ *T*_*Pred*_, the root mean squared error for prediction is

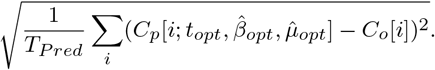

## V. Application on real data

### A. R-package

An R-package has been developed to help the users easily implement the developed methodology with their data. The R-package is available from https://github.com/abh2k/sisd, with detailed instructions for its use. The package is highly flexible in terms of different user-supplied values like *T*_*Current*_, *T*_*Start*_, *T*_*Limit*_ and *T*_*Pred*_ etc. Given the appropriate data and other required input parameter values, the R-package will provide 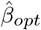 and 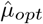, root mean squared error (based on ‘in-sample’ validation), predicted cumulative infected cases.

### B. Predicting cumulative infected COVID-19 cases for Delhi

We consider the COVID-19 data from Delhi, India’s capital city with a population size of around 20 million, to demonstrate the proposed algorithm’s prediction performance. Delhi observed more than 600 thousands of cumulative COVID-19 infected cases at the end of 2020. The data is publicly available from https://www.covid19india.org/.

In Figure 4, we have considered four different *T*_*Current*_ as 29 May 2020 (in (A)), 24 July 2020 (in (B)), 29 December 2020 (in (C)) and 15 January 2021 (in (D)). This set-up can check the proposed algorithm’s prediction performance using the modified SIS model concerning different time periods. Table I shows the 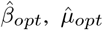, 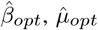, root mean squared error for prediction and other related information. From all the four graphs in Figure 4, it is evident that the proposed algorithm is working well to predict the cumulative infected cases with two different prediction periods 30 (for (A) and (B)) and 40 (for (C) and (D)). From Table I, we see that the chosen optimal training periods’ lengths can be different with different values of 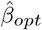, 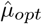. Notice that 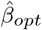 is decreasing over time from 0.12 to 0.09 for Delhi, whereas 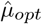 increases for the first three scenarios (from 0.007 to 0.091) then dropped a little to 0.070. These observations support the idea of estimating the modified SIS model’s parameters based on the optimal training phase instead of the entire history as the training phase.

**TABLE I.**
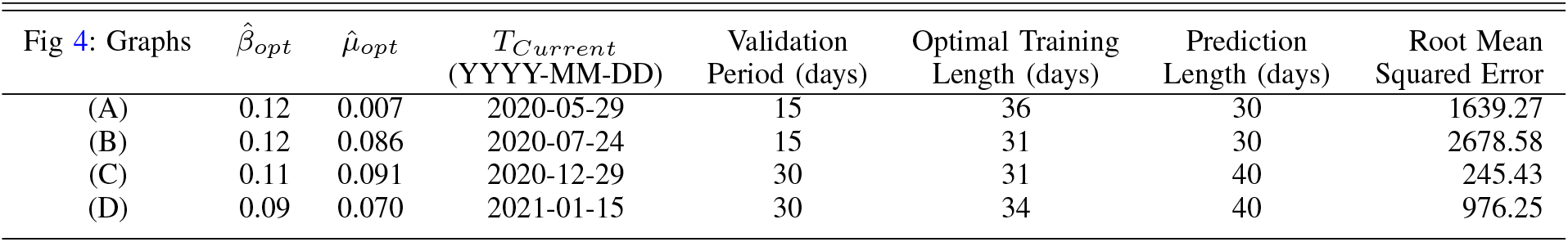
Summary of prediction process for Delhi

| Fig 4: Graphs | $\hat{\beta}_{opt}$ | $\hat{\mu}_{opt}$ | $T_{Current}$<br>(YYYY-MM-DD) | Validation<br>Period (days) | Optimal Training<br>Length (days) | Prediction<br>Length (days) | Root Mean<br>Squared Error |
| --- | --- | --- | --- | --- | --- | --- | --- |
| (A) | 0.12 | 0.007 | 2020-05-29 | 15 | 36 | 30 | 1639.27 |
| (B) | 0.12 | 0.086 | 2020-07-24 | 15 | 31 | 30 | 2678.58 |
| (C) | 0.11 | 0.091 | 2020-12-29 | 30 | 31 | 40 | 245.43 |
| (D) | 0.09 | 0.070 | 2021-01-15 | 30 | 34 | 40 | 976.25 |

**Fig. 4.**
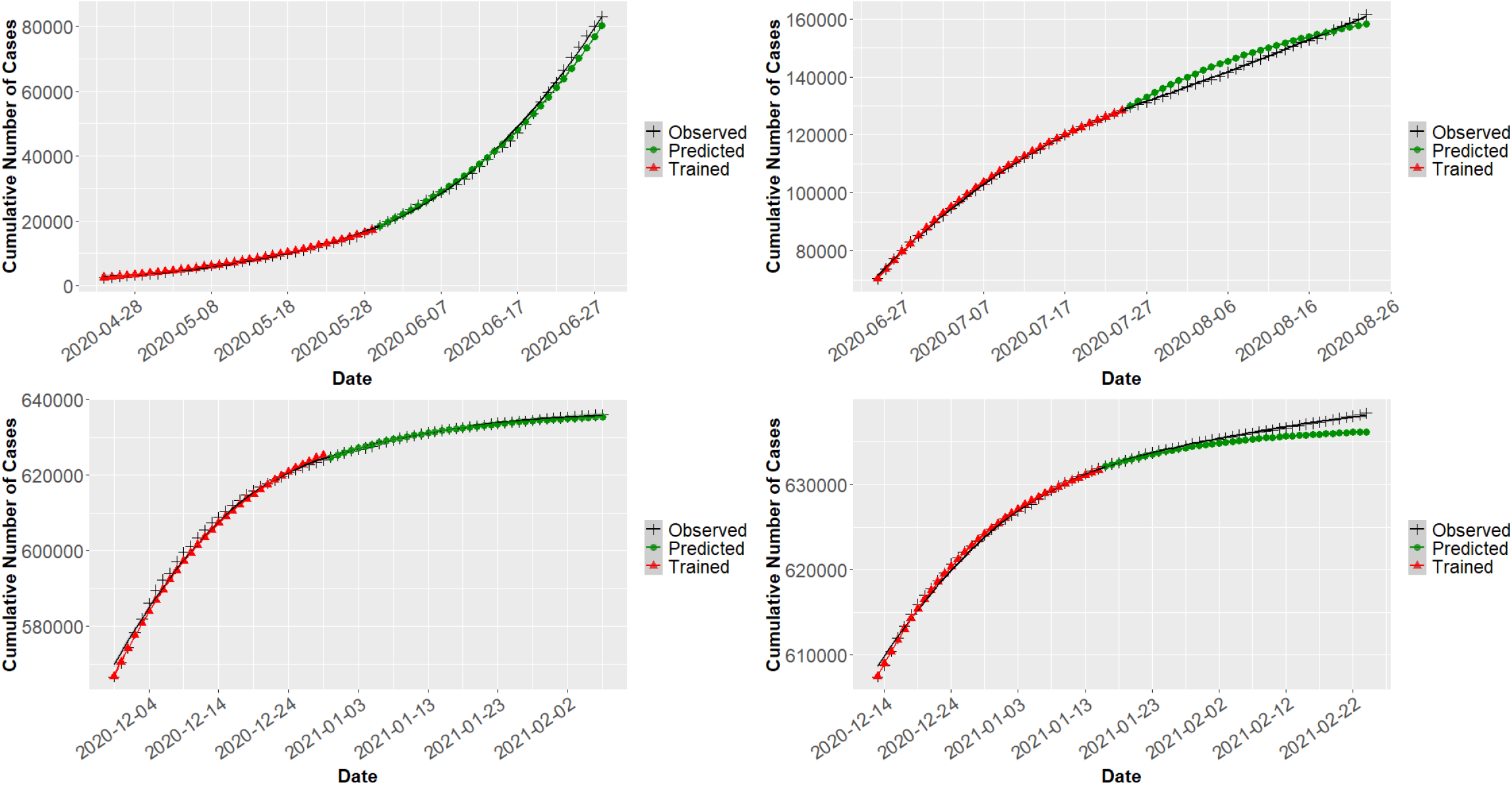
Modified SIS Model predictions based on optimal training phases for Delhi at different periods.

Figure 5 shows what could happen if we include the entire history as a training phase to estimate the model parameters. The 30-day prediction curve based on the entire history (125 days) is exponentially higher than the observed curve of the cumulative infected cases (root mean squared error = 175884.1). The difference between the two curves is getting much bigger for the latter part of the prediction period. However, the prediction curve based on the optimal training phase (total 23 days with 15 days of validation phase) is closer to the curve of observed cumulative infected cases (root mean squared error = 3090.25). The estimated model parameter 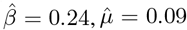 based on the entire history, whereas the same are 0.1 and 0.06 using the optimal training phase, respectively. The 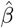 is quite higher in the case of entire history compared to the same 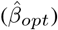 based on the optimal training period. It suggests that the estimation of *β* should be based on an optimal training period to capture the most recent trend rather than the overall trend using entire history.

**Fig. 5.**
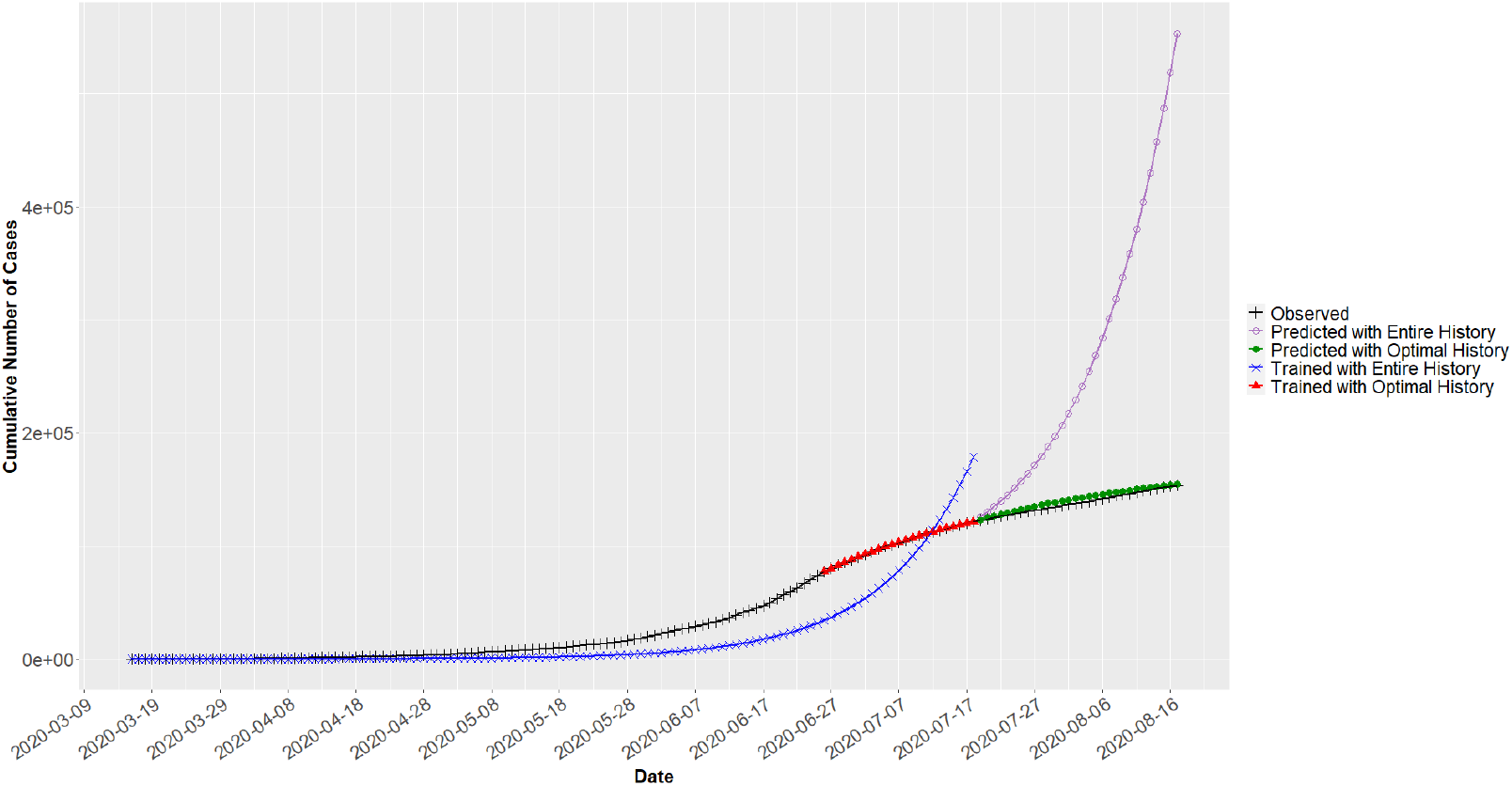
Comparison of the modified SIS Model predictions for Delhi based on the optimal training phase and the entire history period.

### C. Importance of Deceased Compartment

Incorporating the deceased compartment into the modified SIS model is crucial because death due to disease may not be negligible. For example, in COVID-19, the number of deaths to the number of people infected is significant in many countries. Figure 6 shows the importance of the deceased compartment in the modified SIS model in terms of *µ* for Delhi. The prediction curve with 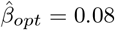 8 and pre-fixed *µ* = 0 (no deaths due to disease) is away from the observed cumulative infected cases, and the difference between the two curves keeps increasing over time, with root mean squared error 12990.46. The prediction curve with 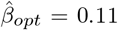 and 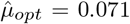 is much closer to the observed cumulative infected cases curve with root mean squared error 5187.47. In the both scenarios, the prediction phase and the validation phase are of 40 and 15 days, respectively.

**Fig. 6.**
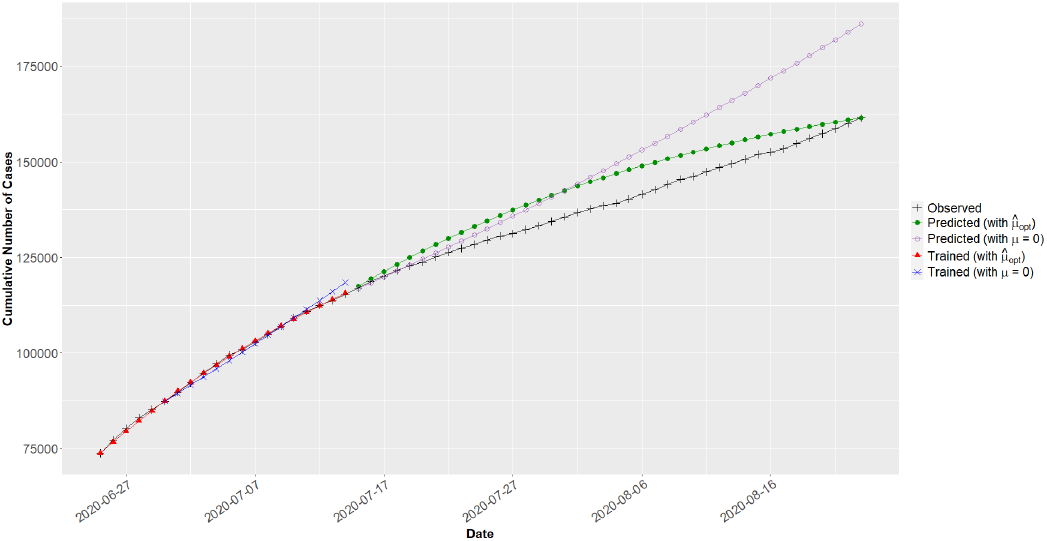
Shows the importance of the deceased compartment (***µ***) in the modified SIS model for better prediction performance.

## VI. Discussion

This work has provided a modified SIS model that accounts for deaths due to disease and predicts cumulative infected cases based on an optimally chosen training phase. The estimation process described in this work is beneficial when the disease under study changes its spreading pattern over time. We have developed the modified SIS model considering COVID-19 as the disease under focus. However, the model and algorithms can be applied to predict the cumulative cases of other infectious diseases.

Even though one can predict for any period-length in the future using the developed model, we recommend restricting the prediction to the short-term only. Any prediction with more than 30 days may not be reliable due to continuous changes in the COVID-19 virus’ characteristics and human behavior (e.g., how social distancing norms followed from time to time).

The developed open-access R-package (https://github.com/abh2k/sisd) can be helpful to implement the modified SIS model without dealing with mathematical details of the model. One only needs to prepare the input data set as described in the R-package documentation.

## Data Availability

We have used open access data. The link to the source data has been mentioned in the manuscript.

https://www.covid19india.org/

## Notes

### Competing Interest Statement

The authors have declared no competing interest.

### Funding Statement

No external funding was received.

